# Large-scale fMRI dataset for the design of motor-based Brain-Computer Interfaces

**DOI:** 10.1101/2024.07.29.24311044

**Authors:** Magnus S Bom, Annette MA Brak, Mathijs Raemaekers, Nick F Ramsey, Mariska J Vansteensel, Mariana P Branco

## Abstract

Functional Magnetic Resonance Imaging (fMRI) data is commonly used to map sensorimotor cortical organization and to localise electrode target sites for implanted Brain-Computer Interfaces (BCIs). Functional data recorded during motor and somatosensory tasks from both adults and children specifically designed to map and localise BCI target areas throughout the lifespan is rare. Here, we describe a large-scale dataset collected from 155 human participants while they performed motor and somatosensory tasks involving the fingers, hands, arms, feet, legs, and mouth region. The dataset includes data from both adults and children (age range: 6-89 years) performing a set of standardized tasks. This dataset is particularly relevant to study developmental patterns in motor representation on the cortical surface and for the design of paediatric motor-based implanted BCIs.

## 1. Background & summary

Brain Computer Interfaces (BCIs) are devices that can convert brain signals to commands to control a computer, providing, for example, severely paralysed individuals with capabilities they have lost due to their paralysis. Specifically, communication BCIs (cBCIs) aim to address severe loss of communication **(Nicolas-Alonso & Gomez-Gil, 2012)**. For implanted cBCIs (e.g., **Vansteensel et al., 2016; Oxley et al., 2020; Metzger et al., 2023; Card et al., 2024; Angrick et al., 2024**) the precise identification of electrode target areas prior to implantation is crucial for optimal performance. Functional magnetic resonance imaging (fMRI) is an effective tool for such identification due to its non-invasive properties, high spatial resolution, and high correlation with BCI performance **(Ramsey et al., 2006; Hermes et al., 2012; Leinders et al., 2023; Piantoni et al., 2021; van den Boom et al., 2021)**.

For BCIs that are controlled by (attempted) movements, signals are usually extracted from the primary sensorimotor cortex (SMC). Common SMC regions for extraction of BCI control signals are the hand, mouth, and foot areas. Many studies have investigated activation patterns in healthy young adults during simple tasks involving hand or finger movement. Such tasks have been found to robustly activate the hand and finger area of the SMC (e.g., **Kleinschmidt et al., 1997**). These findings extend into aging populations (e.g., **Hutchinson, 2002, Ward et al., 2008**), and provide a good basis for motor based BCI use in adults.

Motor based BCIs could also potentially be used to help children with severe communication impairment due to for example Cerebral Palsy. Considerations for implanting BCIs in children is gaining attention in the field of BCIs (e.g., **Kinney-Lang et al., 2016, 2020; Orlandi et al., 2021; Bergeron et al., 2023**). Yet, implementing BCIs in children poses challenges separate from those in adults. For example, little research has been dedicated to identifying developmental changes in primary motor cortex (M1) activation from childhood into adulthood. Some studies showed consistent activation of the contralateral SMC in children (e.g., **Rivkin et al., 2003, Guzzetta et al., 2007**) but there may be differences in activation patterns between age groups that could have implications for the design and longitudinal use of implanted BCIs across the lifespan. For example, implanting electrodes at a site that can drive a BCI during childhood but cannot sustain the use of the BCI into adulthood would require re-insertion of the implanted electrodes at an older age, leading to additional surgical risk.

While large fMRI datasets are available to the research community (e.g., **Crotti et al., 2023**), there are to our knowledge few available datasets that use motor tasks that can be used to map and localise sensorimotor areas for BCI control, and of which the motor output is not used merely as a metric of attention (e.g., button pressing). fMRI datasets are also available for both children (e.g., **Wang et al., 2022**) and adults (e.g., **Peelle et al., 2023**), but they commonly include data from either children or adults, often with different tasks, and thus generally do not allow for comparison between age groups.

Here we present the first large-scale fMRI dataset of 155 children and adults performing a standardized set of motor and somatosensory tasks. The dataset includes a total of 471 runs involving the hand and fingers, tongue, as well as other limbs, such as the arms or legs. By making this dataset publicly available, we hope to promote research on the feasibility of implanted BCIs in children and young adults, allowing researchers to address gaps in the literature related to the representation of brain function in the sensorimotor areas from childhood into adulthood relevant to BCI control in children.

## 2. Methods

### 2.1 Participants

The dataset was obtained from several studies performed at the University Medical Centre Utrecht over the last 15 years. Data was collected from 155 participants (mean age: 35.5±21.3, range: 6-89, 49.7% (78) females, 88.5% (139) right-handed and 1.9% (3) ambidextrous, **Table 1**) who performed 471 tasks in total. Some participants (N = 63) were admitted to the hospital for diagnostic procedures related to their medication-resistant epilepsy (N = 60) or surgical removal of a tumour (N = 3, sub-16, –21 and –38). As part of the pre-surgical workup, these patients underwent fMRI recordings and participated in fMRI sensorimotor experiments for clinical or research purposes. Other participants were healthy volunteers (N = 92) and participated in studies targeting functional mapping of movement. All participants gave written informed consent to participate in the research for which data was acquired and for the use of their data for research purposes. For participants under the age of 18, the informed consent was obtained from the participant’s parents and/or legal guardian. If they were older than 12, these participants also signed an informed consent form. Given that the data is fully anonymous (defaced, randomized and without key) and shared using the BIDS format (https://bids.neuroimaging.io/, **Gorgolewski et al., 2016**), no extra consent was required according to applicable rules and regulations in the Netherlands.

**Table 1.**
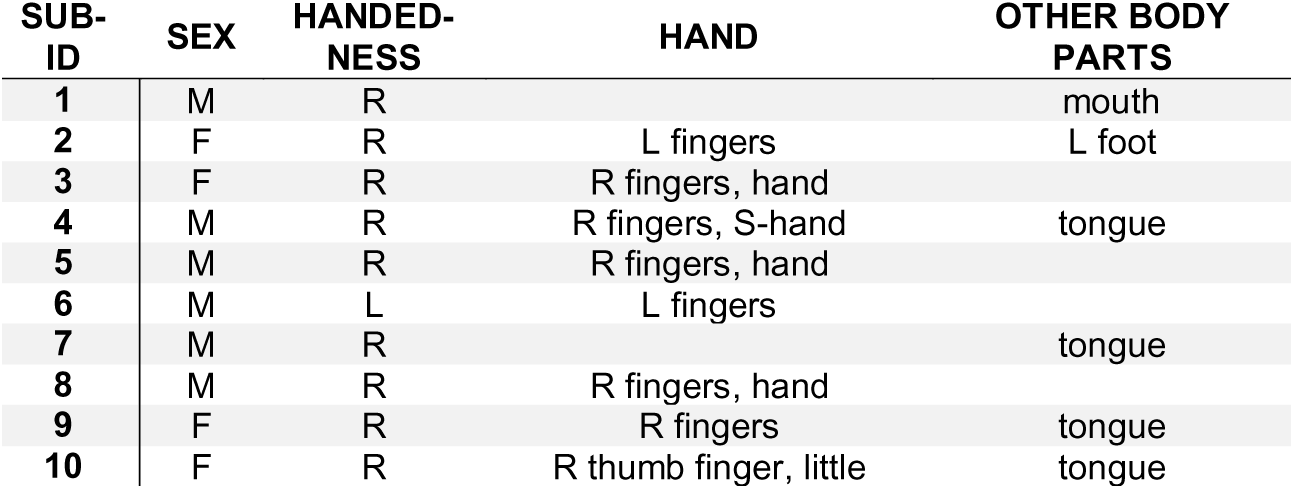
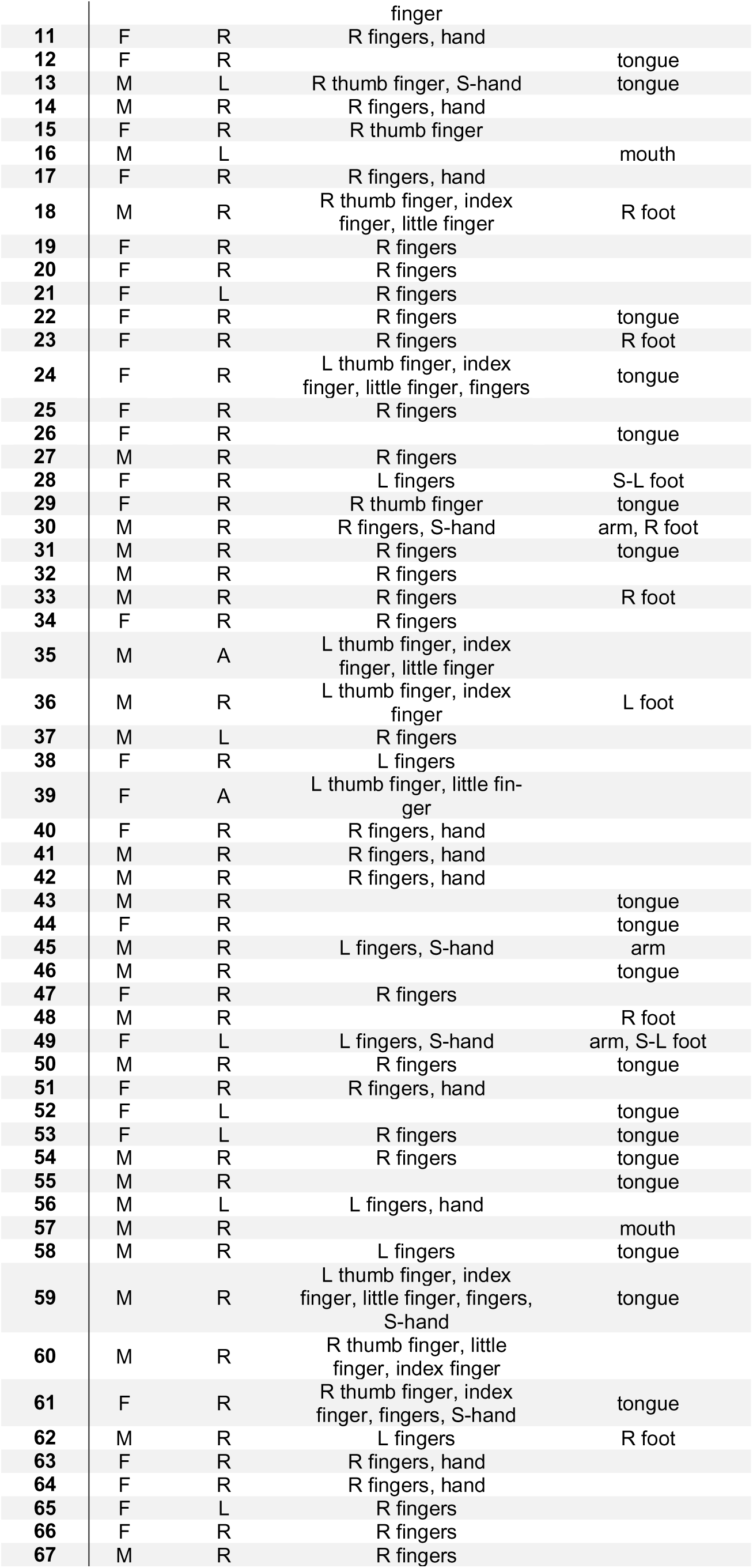
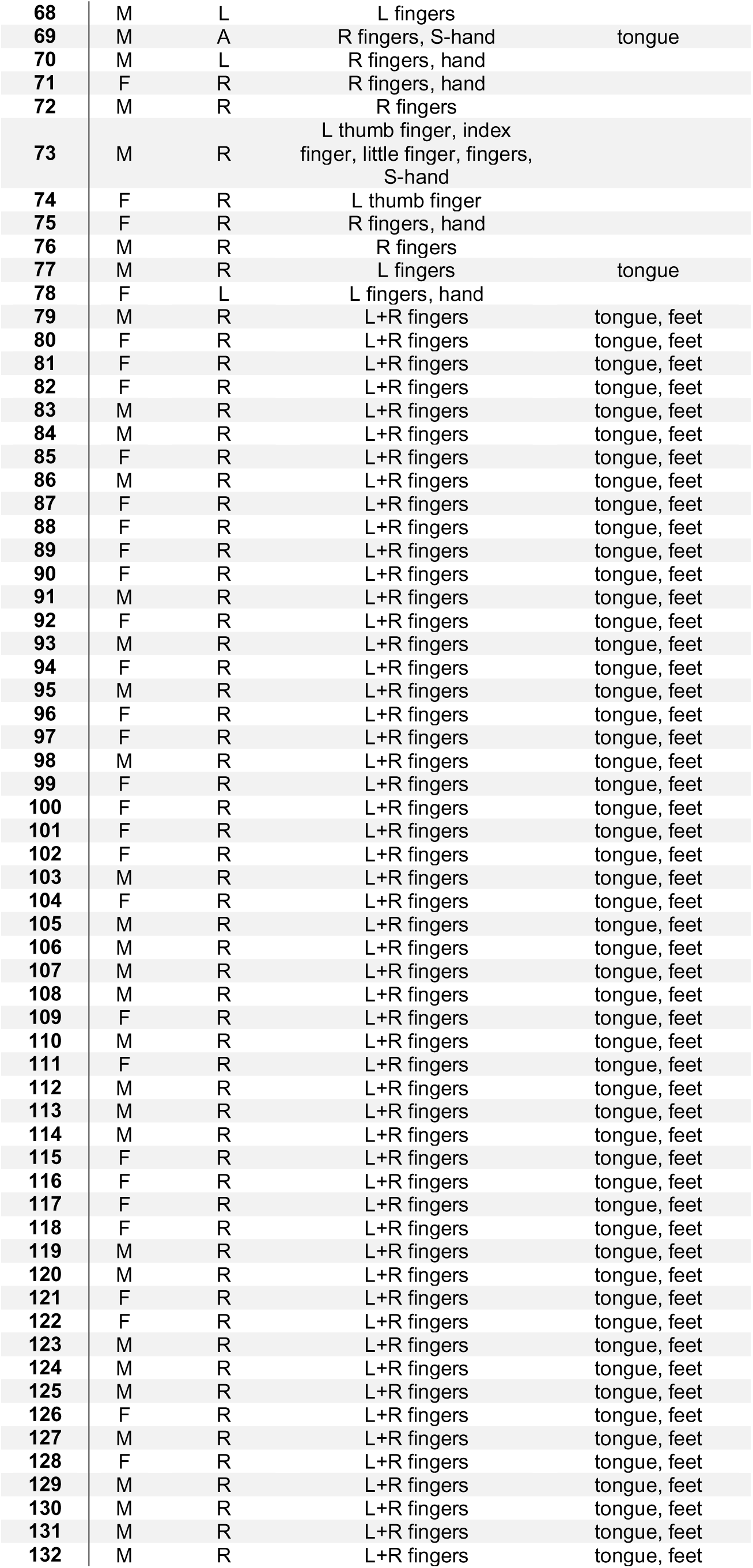
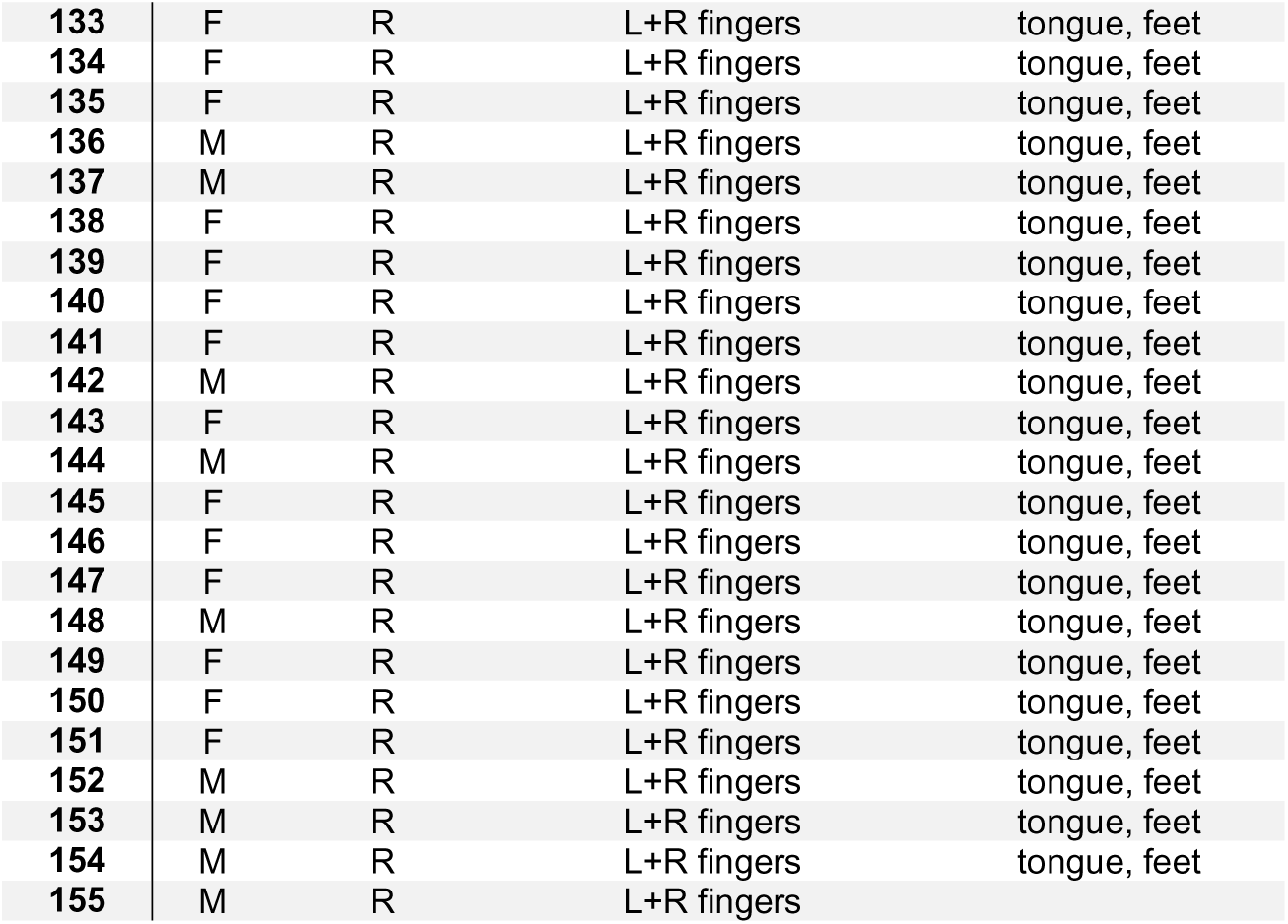
– Demographics. Information about the subject’s ID number, sex (F=female, M=male), handedness (R=right hand, L=left hand), type of movement performed (fingers=fingertapping or movement of multiple individual fingers; hand=open/close hand or hand palm; feet=both feet simultaneously; other body parts), the body side (R=right, L=left; only indicated once if all body parts where performed with the same side), as well as which body part was somatosensory stimulated (S).

### 2.2. Experimental procedures

Data was collected during performance of one of six different standardized tasks specifically designed for sensorimotor localization of hands, mouth or feet **(Figure 1A)**: ‘Motor2Class’, ‘Motor2ClassKids’, ‘Sensory2Class’, ‘Motor3Class’, ‘Mapping3Fingers’, and ‘Mapping5Fingers’. A total of 471 runs were acquired, of which 88.32% (416) consisted of ‘Motor2Class’. All participants completed at least one task, and some participants completed multiple tasks and/or multiple runs of the same task with multiple body parts.

**Figure 1.**
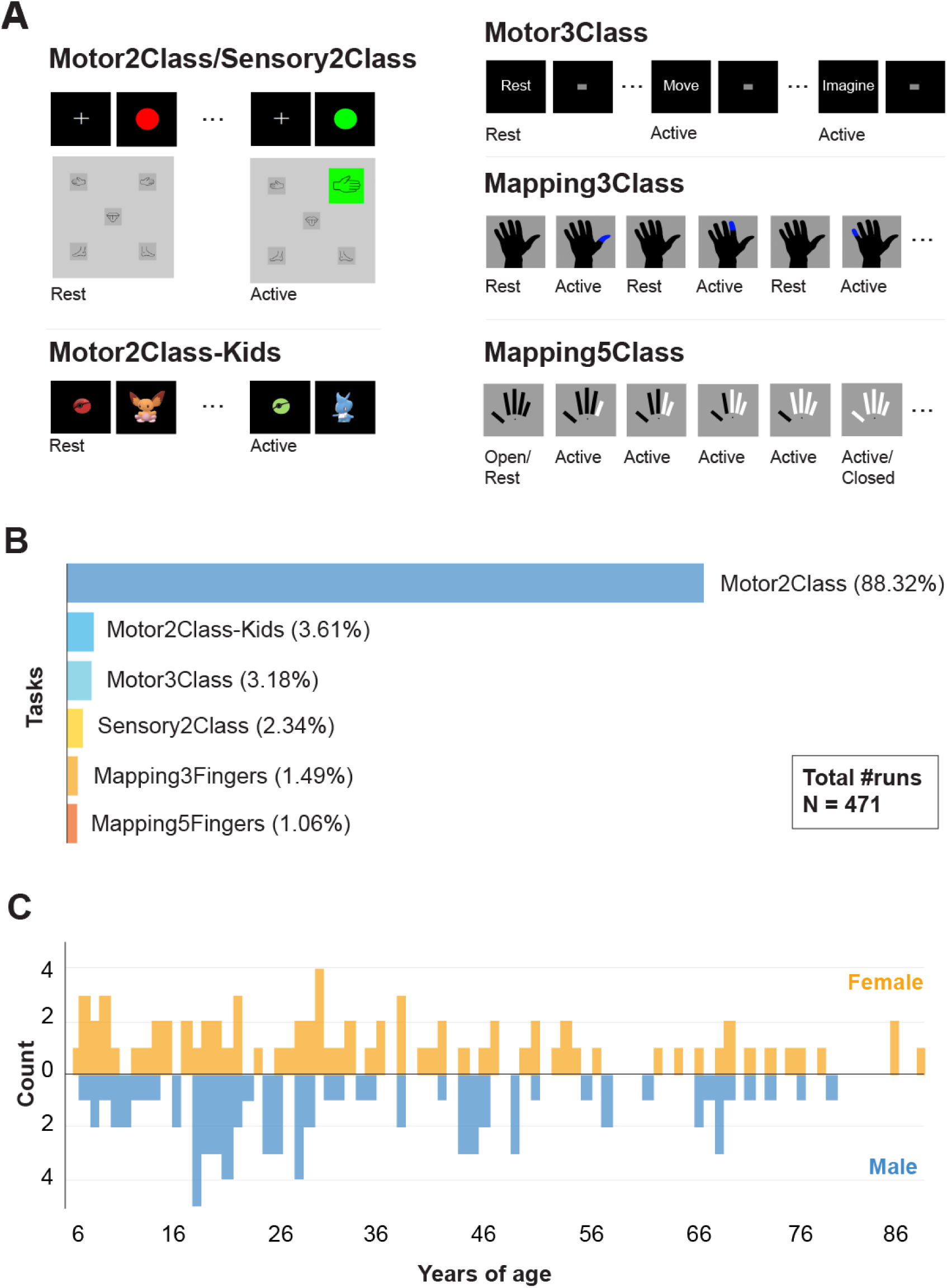
– Overview of tasks and demographics. A) Illustration of task paradigms for Motor/Sensory2Class, Motor2ClassKids, Motor3Class, Mapping3Class and Mapping5Class. A short video of each task can be found in the folder ‘task_examples’. B) Percentage of task-related runs out of total 471 collected runs (across all participants). C) Histogram of age distribution for females (yellow) and males (blue) in the dataset.

#### 2.2.1 Motor2Class and Sensory2Class

The Motor2Class and Sensory2Class were block design tasks with two conditions: rest and active **(Figure 1A)**. During the ‘rest’ condition the participant was asked to lie as still as possible while fixating at the center of a screen. During the ‘active’ condition, the participant was asked to either continuously move a body part (Motor2Class) or to rest while a body part would be stimulated with a brush by a researcher (Sensory2Class). For all Sensory2Class and most Motor2Class tasks, the stimulus presentation included a black screen with blinking circles cuing active (green circle) or rest (red circle) blocks. For the majority of participants, each block lasted for 30s, with a full task consisting of 4 active blocks and 5 rest blocks for 3T and 7T scans, and 8 active blocks and 9 rest blocks for 1.5T scans. For a few Motor2Class scans (sub-9, 66 and 155), each block had varying durations (13-43s) with a full task consisting of 7 active blocks and 8 rest blocks. One participant performed 8 active blocks and 9 rest blocks, with active blocks lasting for 30s. For other participants (sub-79-154, all 3T) an alternative block-design task was used for motor localization, where pictures of body parts were depicted on a screen, in grey during rest blocks. In this design, a full task consisted of 5 active blocks and 6 rest blocks or 7 active blocks and 8 rest blocks of varying durations (8-43s and 7-44s). During active blocks the body part that was requested to move was blinking in green. The body parts included in this dataset using the above tasks were thumb, index or little fingers, all fingers in a ‘fingertapping’ fashion (‘fingers’), hand (open/close movement), arm, feet (both feet simultaneously), lip (kissing movement), tongue (left-right movement) and other general mouth movements **(Table 1)**. The tasks were carried out with either the right or the left body part, except in the case of feet, lip, tongue and mouth movements. Somatosensory stimulation with a brush was performed to whole hand or foot.

#### 2.2.2 Motor2ClassKids

The Motor2ClassKids is child-friendly version of the Motor2Class task **(Figure1A)**. Similarly, the block design task consisted of alternating ‘rest’ and ‘active’ blocks. During the ‘rest’ condition the child was asked lay as still as possible and look at pictures of a cartoon alternating with a picture of a red ‘ball’. During the ‘active’ block, a green ‘ball’ blinked alternating with a ‘cartoon character’ cuing the movement. The participant was instructed to squeeze of a rubber-bulb with the hand every time a ‘cartoon character’ would appear on the screen (body part ‘hand’ in **Table 1**). Each squeeze provided feedback to the participant by displaying a coloured line around the image (i.e., the ‘cartoon character’ was caught with the ‘ball’). A complete run consisted of 5 active blocks and 5 rest blocks (22-30s duration) each containing 11 ‘cartoon characters’ (for more details see **Vansteensel et al., 2021**).

#### 2.2.3 Motor3Class

The Motor3Class task is an alternative block design task with three conditions: one rest and two active conditions, namely overtly/executed movements and imagined movements **(Figure 1A)**. Each condition was cued with an instruction ‘rest’, ‘move’ or ‘imagine’, followed by a block with a blinking grey square. In the ‘rest’ condition the participant was asked to lay as still as possible while fixating on the screen. During the active conditions the participant was asked to continuously move the prespecified body part (see **Table 1**). During the ‘move’ blocks, the participant was required to execute movements overtly, while during the ‘imagine’ blocks the participant imagined performing the same movements. There were a total of 10 blocks per condition, and each block had a duration of 17s including a 1.3s instruction screen (for more details see **Hermes et al., 2011**). The body parts included in the dataset were either movement of all fingers in a ‘fingertapping’ fashion (‘fingers’), foot or tongue **(Table 1)**. The movement or imagination of movement involved the right or the left body part, except for the tongue.

#### 2.2.4 Mapping3Fingers

The Mapping3Fingers task mapped three individual fingers: thumb, index and little fingers using an event-related design **(Figure 1A)**. Each movement consisted of two flexions of the specified finger. A black contour of a hand was displayed on a screen. The movement of the fingers was cued by highlighting the thumb, little finger or index finger in blue for 0.5s. The task consisted of 30 cues of each finger presented in randomized order with an inter-trial interval of 4.4s (for more details see **Siero et al., 2014**).

#### 2.2.5 Mapping5Fingers

The Mapping5Fingers task also used an event-related design to map five individual fingers **(Figure1A)**: thumb, index finger, middle, ring and little finger. In this task flexion and extension of fingers was independently cued, and fingers were cued individually and sequentially. Five rectangles representing each of the five fingers were displayed on a screen. The rectangle turned white to cue finger flexion, and black to cue finger extension. Each finger was kept in the same position (flexed or extended) until the movement of that same finger was cued again. Each finger was cued 8 times during the run at 4.8s intervals, except for the first and last finger which had a longer interval of 14.4s (rest block). Participants started the experiment with an open palm (for more details see **Schellekens et al., 2018**).

### 2.3. Data acquisition details

#### 2.3.1 Structural data acquisition

Structural images of the whole brain were acquired on either a 1.5T ACS-NT Philips scanner (N = 8 participants), a 3T Achieva Philips scanner (N = 139 participants) or a 7T Achieva Philips scanner (N = 13 participants). Some participants had multiple structural scans, one per session. The scanning resolution varied between 0.5 and 2 mm. The specific parameters are available for each scan in the JSON sidecar files and the NIfTI (*.nii) file headers of the dataset.

#### 2.3.2 fMRI data acquisition

Functional images were acquired on either a 1.5T ACS-NT Philips scanner (N = 20 scans), a 3T Achieva Philips scanner (N = 434 scans) or a 7T Achieva Philips scanner (N = 17 scans). A PRESTO scanning sequence was used for the 1.5T scans and 3T scans **(Neggers et al., 2008)**. An EPI scanning sequence was used for the 7T scans. Whole brain scans were acquired for all scans except 7T scans, where a limited field of view was used that included the dorsal part of the brain, extending ventrally to include the hand and finger area. The acquisition time per volume ranged between 0.5 and 4.86 seconds, and the voxel size between 1.384×1.384×1.5 and 4.5×4×4 mm. The specific parameters including the scanning sequence are available for each scan in the JSON (*.json) metadata sidecar files and the NIfTI (*.nii) file headers of the dataset.

## 3. Data records

### 3.1 De-identification and defacing of structural images

All personal identifiable information has been removed from the data. All individual structural images were defaced to comply with the requirements for sharing deidentified medical data. The images were defaced using SPM12 (https://www.fil.ion.ucl.ac.uk/spm/).

### 3.2 Conversion to BIDS structure

Data was standardized using Python (version 3.9) and MATLAB (The MathWorks Inc., 2022) tools. Raw PAR/REC (f)MRI files were converted to NIfTI format using the Nibabel library **(Brett et al., 2020)**. Data was converted to Brain Imaging Data Structure (BIDS) using the function data2bids.m from Fieldtrip Toolbox **(Oostenveld et al., 2011)**. The dataset is therefore organized according to the BIDS format **(Figure 2)**: the root folder contains metadata about the description of the dataset (dataset_description.json), the list of participants along with their demographic details (participants.tsv), a folder with task examples (task_examples), and individual data folders per participant named sub-XXX. The order in which participants were saved in the dataset was randomised (sub-79 to 154 were already randomized before being added to this dataset) and the randomisation key has been deleted, making this dataset fully anonymous. The dataset was validated using https://bids-standard.github.io/bids-validator/.

**Figure 3.**
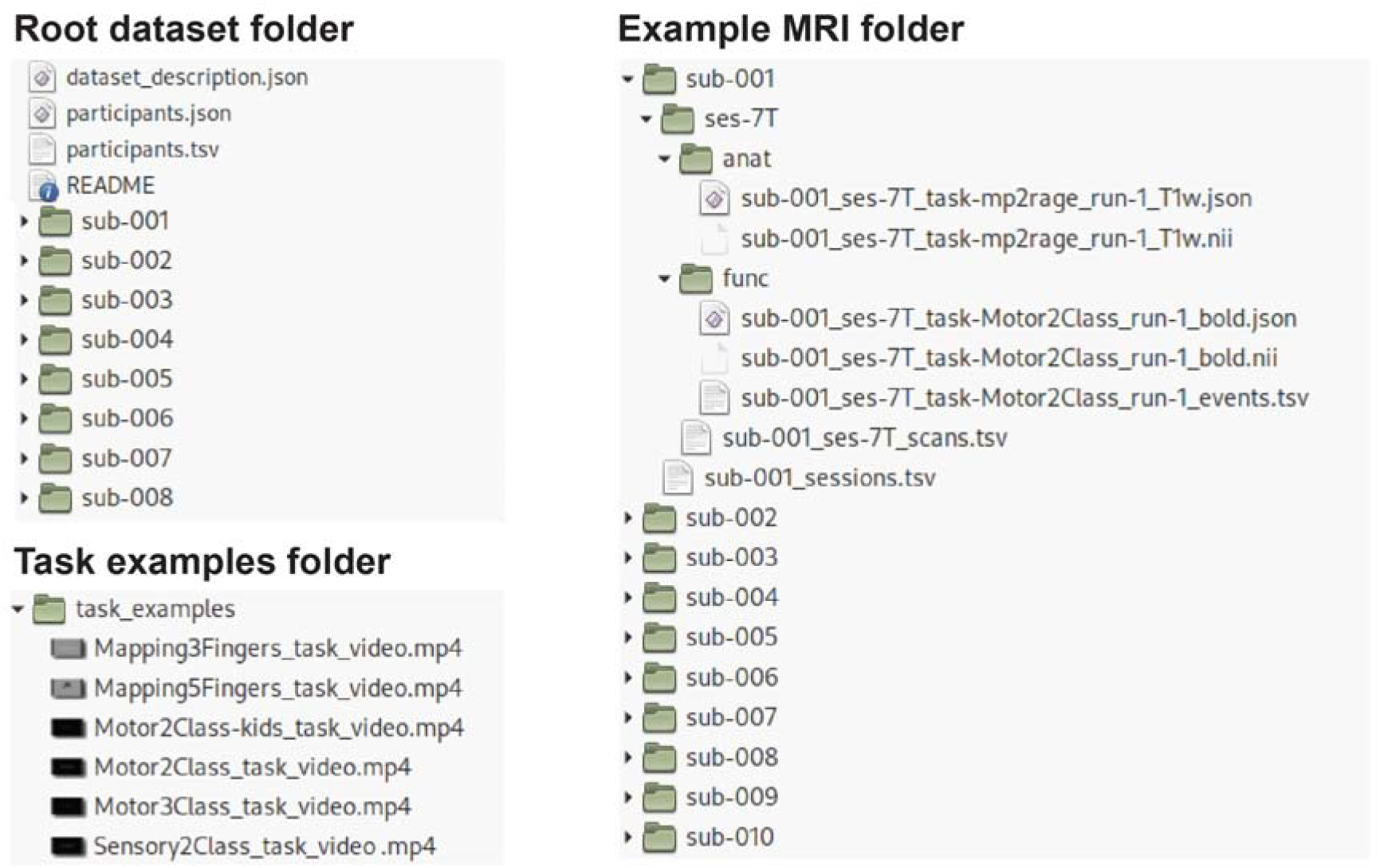
– Overview of BIDS dataset. Structure of files and sub-folders in the root dataset folder and inside subject 001 (sub-001) folder. Each subject folder contains one or more (MRI) sessions with a folder for structural (‘anat’) and functional (‘func’) data. Overview of the task example folder, where short videos of the tasks are saved.

### 3.3 Participant data folders

Each participant’s folder contains one or two sub-directories depending on how many scanning sessions they participated in **(Figure 2)**, typically corresponding to the field strength of the MRI scanner used during the session (ses-1.5T, ses-3T, or ses-7T). Some participants performed two sessions at 1.5T, and so the folders are referred to ses-1.5T and ses-1.5T2, for session 1 and 2 respectively.

### 3.4 (f)MRI folders

Inside each session folder, there are two sub-directories **(Figure 2)**: a folder containing one or multiple anatomical MRI scans (‘anat’), and a folder containing one or multiple functional MRI scans (‘func’). Each participant has an anatomical MRI folder in at least one of their session folders. (f)MRI data are provided in the NIfTI (*.nii) format with sidecar JSON (*.json) files that store additional metadata. Functional images are accompanied by a TSV (*events.tsv) file that contains onsets, event durations (in seconds) and the type of the events of the motor tasks.

### 3.5 Task examples folder

We additionally added a folder containing short videos of each of the five task paradigms **(Figure 2)**. The videos show one of each block type for block designed tasks (Motor2Class, Motor2ClassKids, Sensory2Class and Motor3Class), and roughly 1 minute of the task for event related designs (Mapping3Fingers and Mapping5Fingers). Motor2Class and Sensory2Class share the same video, as they have identical visual stimuli (see **Figure1A**).

## 4. Technical validation

The data was processed with SPM12 and Freesurfer **(Fischl, 2012)**. Preprocessing steps of functional images included realignment, unwarping, and coregistration of the functional images to the anatomical images. Surface reconstructions of the anatomical images were created using the FreeSurfer recon-all pipeline and the functional images were mapped to the ‘fsaverage’ surface while using a smoothing kernel of 6 mm FWHM.

Basic data quality was assessed using a 3-step approach (see details below): 1) we analysed head motion; 2) we correlated activation patterns of individual scans with the mean activation pattern of other participants that performed the same task using the same limb and performed visual inspections of the activation patterns; and 3) we performed visual inspection of the activation patterns of each run. Based on these three metrics we rated each run. Data included in the dataset was deemed good after these three steps as detailed below (**sections 4.1** to **4.3**).

Last, for display purposes we computed the activation pattern in response to the Motor2Class ‘fingers’ task across all participants that performed this task (second-level statistic).

### 4.1 Analysis of head motion

Analysis of head motion involved assessment of the framewise displacement based on the transformation matrices produced by SPM during realignment and unwarping of the raw NIfTI files. The x, y, and z displacement for each voxel and each volume was calculated based on the changes in the transformation matrices following realignment. In addition, motion outliers were calculated independently using FSL (https://fsl.fmrib.ox.ac.uk/) **(Smith et al., 2004; Woolrich et al., 2009)**. This method calculates framewise displacement relative to the first volume and thresholds the displacement in order to classify outliers.

From the included data, the framewise displacement analysis showed roughly 11% of participants had frames with a framewise displacement of more than 4 mm (the largest voxel size in the dataset), while roughly 30% of participants had frames with a framewise displacement of more than 1.5 mm (the smallest voxel size in the dataset) **(Figure 3A)**. In addition, analysis of outliers based on motion showed that 108 participants had more than 5% of their functional volumes marked as outliers **(Figure 3B)**. Increased head motion is expected in fMRI from patient groups and children. Data of young participants may require extra processing steps, such as motion scrubbing **(Power et al., 2014; Charbonnier et al., 2020)**, Volterra expansion for general linear models **(Friston et al., 1996)**, and independent component analysis for artifact removal **(Pruim et al., 2015)**.

**Figure 3.**
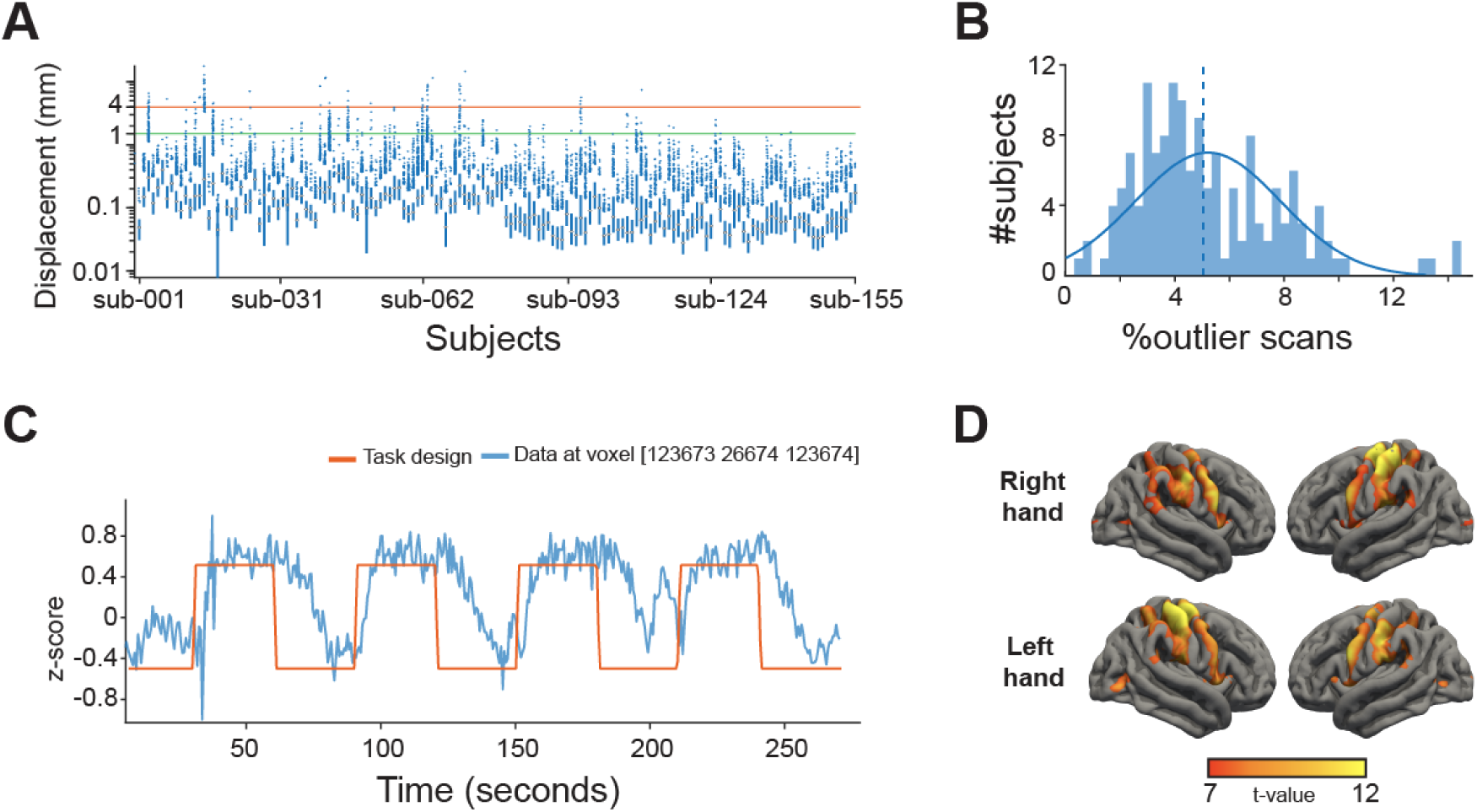
– Technical data validation. A) Boxplot of the framewise displacement metric per subject averaged over scans with maximum and minimum voxel sizes for the dataset marked by green and red horizontal lines, respectively. B) Histogram number of participants with percentage of volumes classified as motion outliers. C) Example time course of the Motor2Class task and observed fMRI activity in one voxel of one subject. D) Second-level group statistics of hand movements from the Motor2Class using ‘fingers’ (fingertapping) for all participants combined, using the t-values on the 90th percentile.

### 4.2 Correlated activation maps

The scans were grouped into four different categories for each hemisphere: hand movements and somatosensory stimulations from block designs, foot movements and somatosensory stimulations from block designs, mouth movements rom block designs, and hand movements from event related designs. For each scan in each group, a mean activation map of all scans from other participants in the same group was calculated **(Jansma et al., 2020)**. The activation map of each scan was then correlated with the mean activation map of the group excluding the current scan. For scans with a correlation with the mean t-map lower than 0.3, the flat surface representation of the t-activation map was inspected. Only scans with task-related activations were kept in the dataset.

### 4.3 Visual inspection of activation maps

Based on the results of the first-level analysis, data were rated based on the level of noise, visually identified as small clusters of t-value activation strewn across the cortex, and the presence and size of clusters of activation in the SMC. Data with fewer clusters and more focal activity in the SMC was deemed good, while data with many dispersed smaller clusters and without an activation in the SMC was deemed bad. All the observed activity patterns in the dataset showed task-related activity and were therefore included in the dataset.

### 4.4 Response to the task

We performed a basic group-level analysis of the response to the task while including all participants. First, a general linear model was fitted to the fMRI data using the block design boxcar function **(Figure 3C)**. Then, we computed a second-level group statistic (in SPM) for the Motor2Class using ‘fingers’ (fingertapping) paradigm, which is the most common task and paradigm in the dataset. Results of analysis overlayed on a surface representation of the ‘fsaverage’ surface showed a clear activation over the left– and right-hand region of the sensorimotor cortex **(Figure 3D)**.

## 5. Usage notes

Under the Public Domain Dedication and License, the data are freely available for non-commercial research purposes. Below we summarise several participant and scan considerations to keep in mind when working with this dataset:

- For hand-related tasks, handedness and the hand used during the experiment did not coincide in 12.1% (N=19) participants.
- The Mapping3Fingers and Mapping5Fingers tasks (participants: 21, 22, 23, 25, 27, 31, 37, 38 and 67), did not include whole-brain scans, which is reflected in the quality metrics. This indicates that special care needs to be taken when dealing with these scans, to ensure that no voxels outside of the scanning area are included in the analysis. Several other participants who performed the Motor2Class (subject 1, 7, 15, 16, 37 and 57) also did not have whole-brain scan but the field-of-view covered most of the sensorimotor cortex.
- The list of participants and scans with more than 10% motion outlier frames for all their scans are listed in **Table 2**.
- The list of participants and scans with a higher framewise displacement than 4 mm (the largest voxel size) for all their scans are listed in **Table 3**.
- For subject 62, session 3T, the “imagine” condition from the Motor3Class task was used for movement of the other hand instead of imagining movement. The movement condition in these files is for right hand movement while the imagine condition is for left hand movement.

**Table 2.**
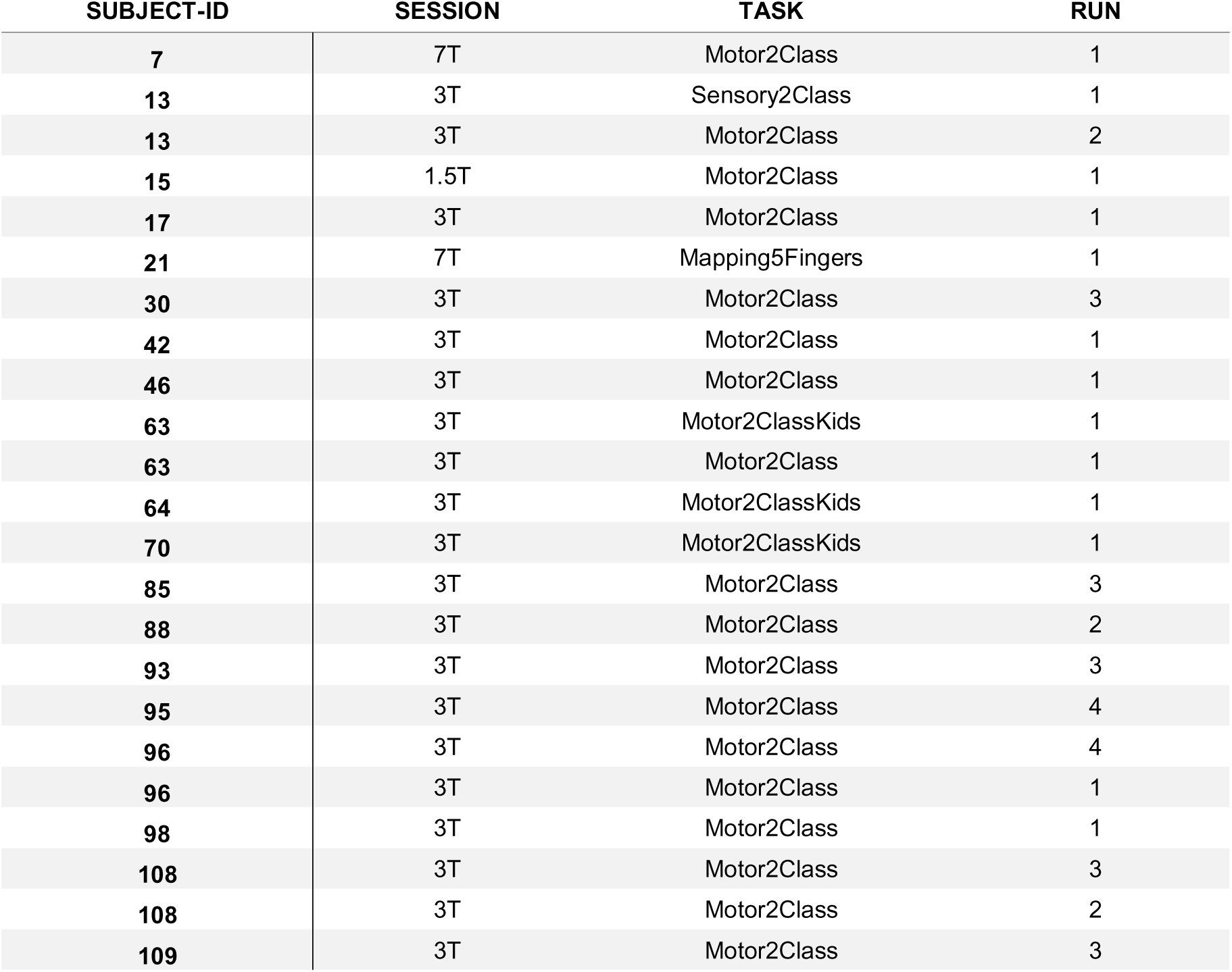
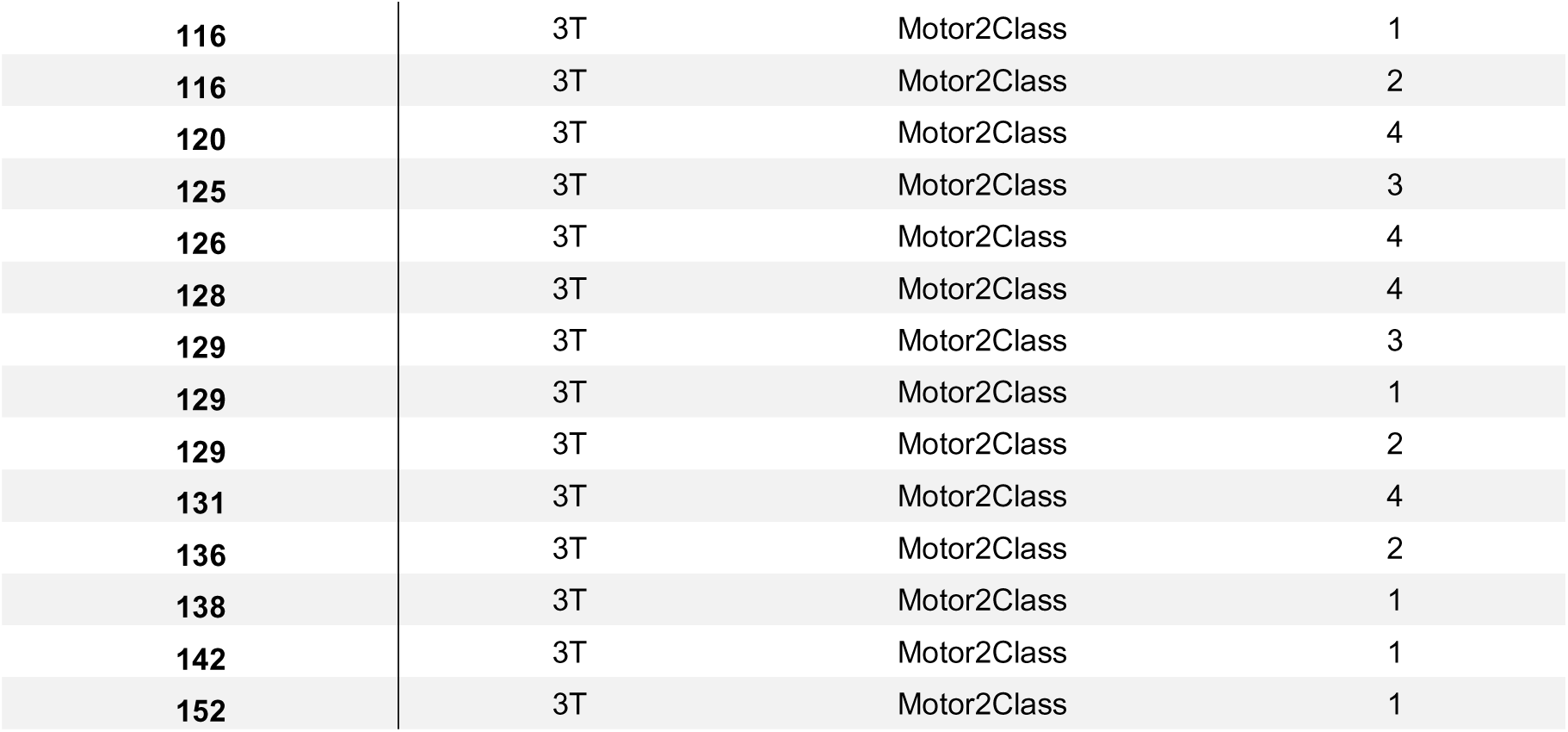
– Participants and scans with more than 10% motion outliers.

**Table 3.** – Participants and scans with framewise displacement larger than 4 mm.

| SUBJECT-ID | SESSION | TASK | RUN |
| --- | --- | --- | --- |
| 3 | 3T | Motor2Class | 1 |
| 3 | 3T | Motor2ClassKids | 1 |
| 7 | 7T | Motor2Class | 1 |
| 13 | 3T | Sensory2Class | 1 |
| 14 | 3T | Motor2Class | 1 |
| 15 | 1.5T | Motor2Class | 1 |
| 17 | 3T | Motor2Class | 1 |
| 25 | 3T | Motor3Class | 1 |
| 40 | 3T | Motor2ClassKids | 1 |
| 42 | 3T | Motor2Class | 1 |
| 46 | 3T | Motor2Class | 1 |
| 51 | 3T | Motor2Class | 1 |
| 62 | 3T | Motor3Class | 1 |
| 63 | 3T | Motor2Class | 1 |
| 63 | 3T | Motor2ClassKids | 1 |
| 70 | 3T | Motor2Class | 1 |
| 70 | 3T | Motor2ClassKids | 1 |
| 71 | 3T | Motor2ClassKids | 1 |
| 96 | 3T | Motor2Class | 4 |
| 109 | 3T | Motor2Class | 4 |

## 6. Code availability

The dataset can be downloaded from the open public repository at https://openneuro.org/datasets/ds005366/. Under the Public Domain Dedication and License, the data are freely available for non-commercial research purposes. The code used to produce the validation metrics are available in the public github repository: https://github.com/UMCU-RIBS/PANDA-fmri-dataset-validation. The ‘main’ function returns several tables containing framewise displacement metrics (function ‘fd_table’) and motion outliers (function ‘fsl_motion_outliers_table’). The functional scans should be aligned and coregistered with the anatomical T1 for each subject before running the ‘fd_table’ function. The function ‘fsl_motion_outliers_table’ requires FSL to be installed and can be used without preprocessing the data. All tables contain four columns that represent the subject number, session, task, and run. Additionally, the framewise displacement table contains an extra fifth column with framewise displacement between the current and preceding frame. Each row represents a frame in each run. The motion outliers table contains 3 additional columns, where the fifth column contains the number of frames, the sixth the number of motion outliers, and the seventh the percentage of frames categorised as motion outliers.

## Data Availability

All data produced are available online at https://openneuro.org/datasets/ds005366/

## Acknowledgments

The authors thank all participants who took the time to participate in the fMRI research conducted in our lab and included in this dataset. This research was supported by the Dutch Research Council (NWO) domain Applied and Engineering Sciences (PANDA project, grant 19072, MPB).

## Contributions

Magnus Bom: data curation, formal analysis and writing original draft. Annette Brak: data curation, formal analysis, review and editing. Mathijs Raemaekers: methodology, review and editing. Nick F Ramsey: resources, review and editing. Mariska Vansteensel: resources, review and editing. Mariana Branco: conceptualization, supervision, funding acquisition, visualization, review and editing.

## Competing interest

The authors declare no competing interests.

## Notes

### Competing Interest Statement

The authors have declared no competing interest.

### Funding Statement

This research was supported by the Dutch Technology Foundation STW (PANDA project, grant 19072)

### Author Declarations

Ethics committee/IRB of UMC Utrecht waived ethical approval for this work because data is fully annonymous.

### Summary of Updates

The content of this manuscript was updated to clarify the methods.

## References

1. Angrick, M., Luo, S., Rabbani, Q., Candrea, D. N., Shah, S., Milsap, G. W., Anderson, W. S., Gordon, C. R., Rosenblatt, K. R., Clawson, L., Tippett, D. C., Maragakis, N., Tenore, F. V., Fifer, M. S., Hermansky, H., Ramsey, N. F., & Crone, N. E. (2024). Online speech synthesis using a chronically implanted brain–computer interface in an individual with ALS. Scientific Reports, 14(1), 9617. 10.1038/s41598-024-60277-2

2. Bergeron, D., Iorio-Morin, C., Bonizzato, M., Lajoie, G., Orr Gaucher, N., Racine, É., & Weil, A. G. (2023). Use of Invasive Brain-Computer Interfaces in Pediatric Neurosurgery: Technical and Ethical Considerations. Journal of Child Neurology, 38(3–4), 223–238. 10.1177/08830738231167736

3. Brett, M., Markiewicz, C. J., Hanke, M., Côté, M.-A., Cipollini, B., McCarthy, P., Jarecka, D., Cheng, C. P., Halchenko, Y. O., Cottaar, M., Larson, E., Ghosh, S., Wassermann, D., Gerhard, S., Lee, G. R., Wang, H.-T., Kastman, E., Kaczmarzyk, J., Guidotti, R., … Freec84. (2020). nipy/nibabel: 3.2.1 (Version 3.2.1) [Computer software]. Zenodo. 10.5281/ZENODO.4295521

4. Card, N. S., Wairagkar, M., Iacobacci, C., Hou, X., Singer-Clark, T., Willett, F. R., Kunz, E. M., Fan, C., Nia, M. V., Deo, D. R., Srinivasan, A., Choi, E. Y., Glasser, M. F., Hochberg, L. R., Henderson, J. M., Shahlaie, K., Stavisky, S. D., & Brandman, D. M. (2024). An Accurate and Rapidly Calibrating Speech Neuroprosthesis. New England Journal of Medicine, 391(7), 609–618. 10.1056/NEJMoa2314132

5. Charbonnier, L., Raemaekers, M. A. H., Cornelisse, P. A., Verwoert, M., Braun, K. P. J., Ramsey, N. F., & Vansteensel, M. J. (2020). A Functional Magnetic Resonance Imaging Approach for Language Laterality Assessment in Young Children. Frontiers in Pediatrics, 8, 587593. 10.3389/fped.2020.587593

6. Crotti, M., Koschutnig, K., & Wriessnegger, S. (2023). *The impact of handedness on the neural correlates during kinesthetic motor imagery: A FMRI study* [Dataset]. Openneuro. 10.18112/OPENNEURO.DS003612.V1.0.3

7. Fischl, B. (2012). FreeSurfer. NeuroImage, 62(2), 774–781. 10.1016/j.neuroimage.2012.01.021

8. Friston, K. J. (2011). Statistical parametric mapping: The analysis of functional brain images. Academic Press.

9. Friston, K. J., Williams, S., Howard, R., Frackowiak, R. S. J., & Turner, R. (1996). Movement-Related effects in fMRI timeseries: Movement Artifacts in fMRI. Magnetic Resonance in Medicine, 35(3), 346–355. 10.1002/mrm.1910350312

10. Gorgolewski, K. J., Auer, T., Calhoun, V. D., Craddock, R. C., Das, S., Duff, E. P., Flandin, G., Ghosh, S. S., Glatard, T., Halchenko, Y. O., Handwerker, D. A., Hanke, M., Keator, D., Li, X., Michael, Z., Maumet, C., Nichols, B. N., Nichols, T. E., Pellman, J., … Poldrack, R. A. (2016). The brain imaging data structure, a format for organizing and describing outputs of neuroimaging experiments. Scientific Data, 3(1), 160044. 10.1038/sdata.2016.44

12. Guzzetta, A., Staudt, M., Petacchi, E., Ehlers, J., Erb, M., Wilke, M., Krägeloh-Mann, I., & Cioni, G. (2007). Brain Representation of Active and Passive Hand Movements in Children. Pediatric Research, 61(4), 485–490. 10.1203/pdr.0b013e3180332c2e

13. Hermes, D., Miller, K. J., Vansteensel, M. J., Aarnoutse, E. J., Leijten, F. S. S., & Ramsey, N. F. (2012). Neurophysiologic correlates of fMRI in human motor cortex. Human Brain Mapping, 33(7), 1689–1699. 10.1002/hbm.21314

14. Hermes, D., Vansteensel, M. J., Albers, A. M., Bleichner, M. G., Benedictus, M. R., Mendez Orellana, C., Aarnoutse, E. J., & Ramsey, N. F. (2011). Functional MRI-based identification of brain areas involved in motor imagery for implantable brain–computer interfaces. Journal of Neural Engineering, 8(2), 025007. 10.1088/1741-2560/8/2/025007

15. Hutchinson, S. (2002). Age-Related Differences in Movement Representation. NeuroImage, 17(4), 1720–1728. 10.1006/nimg.2002.1309

16. Jansma, J. M., Rutten, G.-J., Ramsey, L. E., Snijders, T. J., Bizzi, A., Rosengarth, K., Dodoo-Schittko, F., Hattingen, E., de la Peña, M. J., von Campe, G., Jehna, M., & Ramsey, N. F. (2020). Automatic identification of atypical clinical fMRI results. Neuroradiology, 62(12), 1677–1688. 10.1007/s00234-020-02510-z

17. Kinney-Lang, E., Auyeung, B., & Escudero, J. (2016). Expanding the (kaleido)scope: Exploring current literature trends for translating electroencephalography (EEG) based brain–computer interfaces for motor rehabilitation in children. Journal of Neural Engineering, 13(6), 061002. 10.1088/1741-2560/13/6/061002

18. Kinney-Lang, E., Kelly, D., Floreani, E. D., Jadavji, Z., Rowley, D., Zewdie, E. T., Anaraki, J. R., Bahari, H., Beckers, K., Castelane, K., Crawford, L., House, S., Rauh, C. A., Michaud, A., Mussi, M., Silver, J., Tuck, C., Adams, K., Andersen, J., … Kirton, A. (2020). Advancing Brain-Computer Interface Applications for Severely Disabled Children Through a Multidisciplinary National Network: Summary of the Inaugural Pediatric BCI Canada Meeting. Frontiers in Human Neuroscience, 14, 593883. 10.3389/fnhum.2020.593883

19. Kleinschmidt, A., Nitschke, M. F., & Frahm, J. (1997). Somatotopy in the human motor cortex hand area. A high-resolution functional MRI study. The European Journal of Neuroscience, 9(10), 2178–2186. 10.1111/j.1460-9568.1997.tb01384.x

20. Leinders, S., Vansteensel, M. J., Piantoni, G., Branco, M. P., Freudenburg, Z. V., Gebbink, T. A., Pels, E. G. M., Raemaekers, M. A. H., Schippers, A., Aarnoutse, E. J., & Ramsey, N. F. (2023). Using fMRI to localize target regions for implanted brain-computer interfaces in locked-in syndrome. Clinical Neurophysiology, 155, 1–15. 10.1016/j.clinph.2023.08.003

21. Ma, S., Taicheng Huang, Yukun Qu, Xiayu Chen, Yajie Zhang, & Zonglei Zhen. (2022). *WholebodySomatotopicMapping* [Dataset]. Openneuro. 10.18112/OPENNEURO.DS004044.V2.0.3

22. Metzger, S. L., Littlejohn, K. T., Silva, A. B., Moses, D. A., Seaton, M. P., Wang, R., Dougherty, M. E., Liu, J. R., Wu, P., Berger, M. A., Zhuravleva, I., Tu-Chan, A., Ganguly, K., Anumanchipalli, G. K., & Chang, E. F. (2023). A highperformance neuroprosthesis for speech decoding and avatar control. Nature, 620(7976), 1037–1046. 10.1038/s41586-023-06443-4

23. Neggers, S. F. W., Hermans, E. J., & Ramsey, N. F. (2008). Enhanced sensitivity with fast three-dimensional blood-oxygenlevel-dependent functional MRI: Comparison of SENSE–PRESTO and 2D-EPI at 3 T. NMR in Biomedicine, 21(7), 663–676. 10.1002/nbm.1235

24. Nicolas-Alonso, L. F., & Gomez-Gil, J. (2012). Brain Computer Interfaces, a Review. Sensors, 12(2), 1211–1279. 10.3390/s120201211

25. Oostenveld, R., Fries, P., Maris, E., & Schoffelen, J.-M. (2011). FieldTrip: Open Source Software for Advanced Analysis of MEG, EEG, and Invasive Electrophysiological Data. Computational Intelligence and Neuroscience, 2011, 1–9. 10.1155/2011/156869

26. Orlandi, S., House, S. C., Karlsson, P., Saab, R., & Chau, T. (2021). Brain-Computer Interfaces for Children With Complex Communication Needs and Limited Mobility: A Systematic Review. Frontiers in Human Neuroscience, 15, 643294. 10.3389/fnhum.2021.643294

27. Oxley, T., Mitchell, P., Opie, N., Yoo, P., Mocco, J., Campbell, B., Bird, C., & Lee, S. (2020). Motor neuroprosthesis implanted using cerebral venography improves activities of daily living in severe paralysis. J. Neurointervent. Surg., 12, A165– A166.

28. Peelle JE, Spehar B, MS, J., McConkey S, Myerson J, S, H., Sommers MS, & Tye-Murray N. (2023). *Visual and audiovisual speech perception associated with increased functional connectivity between sensory and motor regions* [Dataset]. Openneuro. 10.18112/OPENNEURO.DS003717.V1.1.0

29. Piantoni, G., Hermes, D., Ramsey, N., & Petridou, N. (2021). Size of the spatial correlation between ECoG and fMRI activity. NeuroImage, 242, 118459. 10.1016/j.neuroimage.2021.118459

30. Power, J. D., Mitra, A., Laumann, T. O., Snyder, A. Z., Schlaggar, B. L., & Petersen, S. E. (2014). Methods to detect, characterize, and remove motion artifact in resting state fMRI. NeuroImage, 84, 320–341. 10.1016/j.neuroimage.2013.08.048

31. Pruim, R. H. R., Mennes, M., Van Rooij, D., Llera, A., Buitelaar, J. K., & Beckmann, C. F. (2015). ICA-AROMA: A robust ICA-based strategy for removing motion artifacts from fMRI data. NeuroImage, 112, 267–277. 10.1016/j.neuroimage.2015.02.064

32. Ramsey, N. F., van de Heuvel, M. P., Kho, K. H., & Leijten, F. S. S. (2006). Towards human BCI applications based on cognitive brain systems: An investigation of neural signals recorded from the dorsolateral prefrontal cortex. IEEE Transactions on Neural Systems and Rehabilitation Engineering, 14(2), 214–217. IEEE Transactions on Neural Systems and Rehabilitation Engineering. 10.1109/TNSRE.2006.875582

33. Rivkin, M. J., Vajapeyam, S., Hutton, C., Weiler, M. L., Hall, E. K., Wolraich, D. A., Yoo, S. S., Mulkern, R. V., Forbes, P. W., Wolff, P. H., & Waber, D. P. (2003). A functional magnetic resonance imaging study of paced finger tapping in children. Pediatric Neurology, 28(2), 89–95. 10.1016/S0887-8994(02)00492-7

34. Schellekens, W., Petridou, N., & Ramsey, N. F. (2018). Detailed somatotopy in primary motor and somatosensory cortex revealed by Gaussian population receptive fields. NeuroImage, 179, 337–347. 10.1016/j.neuroimage.2018.06.062

35. Siero, J. C. W., Hermes, D., Hoogduin, H., Luijten, P. R., Ramsey, N. F., & Petridou, N. (2014). BOLD matches neuronal activity at the mm scale: A combined 7T fMRI and ECoG study in human sensorimotor cortex. NeuroImage, 101, 177–184. 10.1016/j.neuroimage.2014.07.002

36. Smith, S. M., Jenkinson, M., Woolrich, M. W., Beckmann, C. F., Behrens, T. E. J., Johansen-Berg, H., Bannister, P. R., De Luca, M., Drobnjak, I., Flitney, D. E., Niazy, R. K., Saunders, J., Vickers, J., Zhang, Y., De Stefano, N., Brady, J. M., & Matthews, P. M. (2004). Advances in functional and structural MR image analysis and implementation as FSL. NeuroImage, 23, S208–S219. 10.1016/j.neuroimage.2004.07.051

37. van den Boom, M. A., Miller, K. J., Ramsey, N. F., & Hermes, D. (2021). Functional MRI based simulations of ECoG grid configurations for optimal measurement of spatially distributed hand-gesture information. Journal of Neural Engineering, 18(2). 10.1088/1741-2552/abda0d

38. Vansteensel, M. J., Pels, E. G. M., Bleichner, M. G., Branco, M. P., Denison, T., Freudenburg, Z. V., Gosselaar, P., Leinders, S., Ottens, T. H., Van Den Boom, M. A., Van Rijen, P. C., Aarnoutse, E. J., & Ramsey, N. F. (2016). Fully Implanted Brain–Computer Interface in a Locked-In Patient with ALS. New England Journal of Medicine, 375(21), 2060–2066. 10.1056/NEJMoa1608085

39. Vansteensel, M. J., Selten, I. S., Charbonnier, L., Berezutskaya, J., Raemaekers, M. A. H., Ramsey, N. F., & Wijnen, F. (2021). Reduced brain activation during spoken language processing in children with developmental language disorder and children with 22q11.2 deletion syndrome. Neuropsychologia, 158, 107907. 10.1016/j.neuropsychologia.2021.107907

40. Wang, J., Lytle, M. N., Weiss, Y., Yamasaki, B. L., & Booth, J. R. (2022). A longitudinal neuroimaging dataset on language processing in children ages 5, 7, and 9 years old. Scientific Data, 9(1), 4. 10.1038/s41597-021-01106-3

41. Ward, N. S., Swayne, O. B. C., & Newton, J. M. (2008). Age-dependent changes in the neural correlates of force modulation: An fMRI study. Neurobiology of Aging, 29(9), 1434–1446. 10.1016/j.neurobiolaging.2007.04.017

42. Woolrich, M. W., Jbabdi, S., Patenaude, B., Chappell, M., Makni, S., Behrens, T., Beckmann, C., Jenkinson, M., & Smith, S. M. (2009). Bayesian analysis of neuroimaging data in FSL. NeuroImage, *45*(1 Suppl), S173–186. 10.1016/j.neuroimage.2008.10.055

